# Dynamic excitation/inhibition balance preceding seizure onset and its link to functional and structural brain architecture

**DOI:** 10.1101/2025.06.16.25329680

**Authors:** Gian Marco Duma, Simone Cuozzo, Alberto Danieli, Justine Y. Hansen, Lisa Antoniazzi, Elisa Osanni, Valerio Vitale, Paolo Bonanni, Giovanni Pellegrino

## Abstract

Altered excitation-inhibition (E/I) balance is a hallmark of epilepsy, yet its dynamic evolution before seizure onset remains unclear. In this study, we investigated time-resolved changes in cortical E/I balance and their network-level and structural correlates in patients with drug-resistant focal epilepsy. High-density EEG recordings encompassing at least one recorded seizure were obtained from pre-surgical evaluations. Using source-reconstructed EEG, we tracked dynamic changes in the aperiodic exponent of the power spectrum—a non-invasive proxy for E/I balance—during preictal and interictal periods. Directed functional connectivity was assessed using spectral Granger causality, and correlations with cortical thickness and neurotransmitter receptor density were evaluated. We found that the aperiodic exponent increased progressively in the minutes preceding seizures, reflecting a widespread shift toward cortical inhibition. This pattern was specific to the preictal state and evident across both epileptogenic and non-epileptogenic regions. Periodic activity in delta and theta bands also increased prior to seizures, supporting a global reorganization of cortical excitability. At the functional profile level, epileptogenic regions demonstrated significantly greater outward connectivity than both their inward connectivity and that of non-epileptogenic areas during the preictal phase. Moreover, E/I balance was differentially related to connectivity patterns: non-epileptogenic regions showed a positive correlation between inhibition (higher aperiodic exponent) and outward connectivity, whereas epileptogenic regions displayed a trend toward the opposite pattern. Structure-function coupling also diverged: cortical thickness positively correlated with inhibitory tone in non-epileptogenic regions but not in epileptogenic ones. Finally, we observed that lower values of muscarinic receptor density corresponded to an E/I balance shifted towards inhibition (ie., higher aperiodic exponent), linking cholinergic tone with preictal E/I dynamics. These findings indicate that seizure onset is preceded by a shift toward inhibition at the whole-brain level, accompanied by distinct patterns of directed connectivity and disrupted structural coupling in epileptogenic networks. This multiscale perspective highlights the interplay between electrophysiology, network topology and cortical architecture systems in the moments leading up to seizure onset. Our results support and enlarge the interictal suppression hypothesis, suggesting a network protective mechanism through inhibition, related to a perturbatory event as the seizure onset. Overall, the dynamic aperiodic exponent emerges as a promising non-invasive marker with potential applications in seizure prediction and network-targeted neuromodulation.

## Introduction

The healthy brain optimizes the balance between excitatory and inhibitory (E/I) signals, resulting in whole-brain functional networks that support the efficient transfer of information across the cortex ^1–4^. Disruptions to this balance represent one of the leading mechanisms in several neurological disorders ^5–9^. Epilepsy, a condition characterized by recurrent seizures, is closely associated with E/I imbalance in which alterations of E/I dynamics facilitate the synchronized firing of neurons across brain networks, ultimately resulting in seizure activity^10–12^.

Over the past decades, several methods have been developed to non-invasively estimate the E/I balance using electrophysiological and neuroimaging data ^13^. The exponent of the aperiodic component of the electroencephalography (EEG) power spectrum captures the broadband, non-oscillatory neural dynamics and has been shown to be a sensitive proxy for the E/I balance in cortical networks ^14^. A steeper (higher or more negative) exponent indicates greater inhibitory activity, whereas a flatter (lower or less negative) exponent suggests increased excitation. This measure potentially allows a non-invasive full-cortex assessment of E/I balance in neurological disorders ^5,15,16^.

A recent study examining the relationship between the aperiodic exponent and epilepsy has demonstrated greater inhibition (i.e., higher values of aperiodic exponent) in epileptogenic regions in the interictal phase (resting state activity), highlighting the relationship of E/I balance with pharmacological treatment and cognitive functioning ^17^.

Additional evidence from our group showed that interictal spikes are accompanied by a surge of inhibitory activity in regions distant from the epileptic focus. This widespread inhibition appears to have a protective role against the disruptive effects of epileptiform activity and is associated with better cognitive outcomes in patients with drug-resistant epilepsy. ^18^. These results align well with the idea that epilepsy is a disorder linked to the imbalance of E/I and with the theoretical framework of the “interictal suppression hypothesis” (ISH). The ISH posits that the epileptogenic regions are characterized by a larger amount of inward functional connections as compared to the rest of the brain. These connections are theorized to be inhibitory inputs representing a protective mechanism isolating the perturbing nodes— namely, the epileptogenic regions—from the rest of the brain ^19,20^. However, the majority of the studies supporting this framework have been performed during the interictal phase, and little is known about the dynamic of the E/I in the transition between interictal and ictal activity.

The transition from interictal to ictal and seizure generation are dynamic processes in which slow variations of global E/I push the system towards a bifurcation and a successive change in the brain state ^21,22^. Recent computational and empirical studies suggest that changes in E/I balance can systematically affect network organization, either enhancing or dampening the influence one region exerts over another ^23^. Therefore, the evolution of the E/I balance and its relationship with effective connectivity on a whole-brain scale may play a crucial role in seizure initiation ^24^. In this light, characterizing E/I dynamics and its relationship to the brain’s functional organization is critical for interpreting neural mechanisms underlying seizure generation, in a network-disorder theoretical framework.

To this purpose, we investigated the dynamics of E/I balance and the directionality of inter-regional functional interactions (i.e., inward and outward connectivity) in the preictal phase (minutes before seizures), using dynamic measurements of aperiodic exponent and spectral granger causality, respectively. We applied this framework to a cohort of drug-resistant epilepsy patients who underwent high-density EEG and cortical source reconstruction of electrical activity of interictal and interictal-ictal transition ^25^.

We hypothesized that the slope of the PSD aperiodic component would increase linearly as seizure onset approached, thereby reflecting a dynamic change in E/I balance leading to the ictal event ^26^, as the seizure approached, suggesting a dynamic change of the E/I balance leading to the ictal event. We also expect that dynamic E/I changes preceding seizures would have downstream effects on the brain’s functional architecture.

E/I balance and large-scale neural dynamics are shaped by local properties such as cortical structure, gene expression, and neurotransmitter receptor densities ^27–29^. Recent findings demonstrate a specific link between the aperiodic EEG activity and cortical expression of genes regulating E/I balance, as well as structural network organization ^17,30^. Investigating structure-function coupling may shed light on the pathophysiological mechanisms of epilepsy, from micro-to macro-scale. To this purpose we studied the influence of cortical thickness and receptor density of neurotransmitters on the E/I in the preictal phase. Overall, the present study aims to better elucidate large scale brain organization in relation to seizure ignition, leveraging non-invasive techniques such as high density EEG.

## Methods

### Participants

Patients with drug resistant focal epilepsy underwent prolonged high density EEG for clinical pre-surgical evaluation between 2021 and 2024 at the Epilepsy and Clinical Neurophysiology Unit, IRCCS Eugenio Medea in Conegliano (Italy). We included patients with at least one seizure recorded with high density EEG. The presurgical workflow included Video EEG monitoring (32 channels, 4 days), 24 hours with high density EEG, brain magnetic resonance imaging (MRI), and positron emission tomography as an adjunctive investigation in selected patients. Twenty patients were retrospectively screened and included in the sample [mean age = 35.50 (SD = 16.94); 9 females; 8 TLE; 6 frontal epilepsy; 4 parietal epilepsy; 2 unknown]. 85% of patients were drug-resistant. Patients’ demographic and clinical characteristics are provided in Table 1. The study protocol was conducted according to the Declaration of Helsinki and approved by the local ethical committee. All participants provided written informed consent to participate in the study.

**Table 1.** Sample demographic and clinical information. Demographic and clinical features of the patients. MRI findings are reported based on neuroradiological examination. Abbreviation of MRI abnormalities: FCD, focal cortical dysplasia; HS, hippocampal sclerosis.

| **Patients** | **Mean ± Standard deviation** |
| --- | --- |
| Age | 35.50 ± 16.94 |
| Age of onset | 20.80 ± 16.65 |
| Number of antiseizure medications | 2.05 ± 1.16 |
| Duration of epilepsy (years) | 14.73 ± 14.90 |
| **MRI** | **Number** |
| Cavernous Angioma | 1 |
| Cerebellar Atrophy | 1 |
| FCD | 9 |
| HS | 3 |
| Polymicrogyria | 1 |
| MRI-negative | 4 |
| Unknown | 1 |

### EEG recording and preprocessing

High density EEG was recorded with a 128-channel Micromed System Plus device. The signal was sampled at 512 Hz and referenced to the vertex. The impedance of the electrodes was kept <5 kΩ.

Seizure onset was marked by two epileptologists with expertise in EEG (AD and PB). For each subject we collected 10 minutes of inter-ictal (resting state) hdHDEEG activity with a separate recording with an average distance from the seizure of 3.2 ± 1.25 (standard deviation; DV) hours.

Signal preprocessing was conducted using EEGLAB version 14.1.2b44 ^31^, following a pipeline validated in previous work ^32,32^. The preprocessing steps included: (a) data resampling to 250 Hz; (b) bandpass filter (0.1–45 Hz) with Hamming windowed sinc finite impulse response filter; (c) 1-second data epoching; (d) rejection of epochs and channels with artifacts; (e) average re-reference; (f) bad channels interpolation (spherical spline method); (g) independent component analysis (ICA) (40 components-Infomax algorithm); (h) automatic artifact-related ICA components removal and (i) concatenating the epochs to reconstruct a continuous signal.

The automatic artifact and bad-channel detection was performed using the Trial-by-Trial plugin in EEGLAB with the following parameters: channels were marked as bad if their differential average amplitude exceeded 250 μV in more than 30% of the epochs. Epochs were excluded if they contained more than 10 bad channels. Flat channels were detected and removed using the Trimoutlier plugin, with a lower threshold of 1 μV. For the automatic ICA rejection, we used the ICLabel plug-in ^33^. The ICA components classified as brain were kept (after visual inspection), while components classified as eyes, muscle, channel or line noise, heart or others within a 70-100% accuracy range were removed.

### Data

Duration of preictal EEG data ranged from 2 to 160 minutes. On average, 2.93 ± 3.75 (standard deviation; DV) channels were interpolated, 432.80 ± 504.21 epochs and 12.48 ± 4.66 components were rejected. After excluding two patients for whom preictal data lasted less than 5 minutes, the minimum preictal data duration after the cleaning was 13 minutes. In order to be consistent across participants, we selected for each subject 13 minutes of signal prior to the seizure. For the interictal activity, on average, 0.11± 0.48 channels, 44.64 ± 9.51 epochs and 12.35 ± 4.60 components were rejected.

After preprocessing, each subject had at least 7.5 min of artifact-free continuous signal. To harmonize data across subjects, we selected 450 seconds for the interictal (resting state) data of each participant.

### Cortical source modelling

Individual MRIs used for source imaging consisted of high-resolution T1-weighted three-dimensional (3D) isotropic (1mm) acquisitions, collected at Ospedale San Bortolo, Vicenza, with a Siemens Skyra scanner 3T. The images were segmented using the Freesurfer (7.3.2) standard pipeline with default parameters ^34,35^. Brain surfaces and Freesurfer-derived cortical thickness maps were then imported into Brainstorm ^36^.

Electrode co-registration with MRI anatomy was carried out in Brainstorm using standard anatomical landmarks, with manual adjustments applied if necessary. The cortical surface mesh was downsampled to 15,002 vertices. Three boundary element model (BEM) surfaces—inner skull, outer skull, and scalp—were reconstructed (1082 vertices per surface). The forward model was computed using a three-shell BEM approach (default Brainstorm parameters) using Brainstorm integrated OpenMEEG plugin ^37,38^.

For the inverse solution, we employed the weighted minimum norm estimate, using Brainstorm’s default parameter ^39^ settings. Source activity was then downsampled to 68 cortical regions of interest (ROIs) according to the Desikan-Killiany ^40^ atlas. ROI time series were obtained as the mean time series across ROI vertices ^41^. The same atlas mask was applied to cortical thickness maps to obtain a thickness value per ROI. Finally, for each subject we identified the Epileptogenic Regions as the cortical parcel containing the epileptogenic focus extracted from clinical reports. By contrast, all remaining cortical parcels were classified as Non-Epileptogenic regions.

### Dynamic parameterization of aperiodic and periodic activity

Source space time-series from ROIs were processed with the SPRINT Brainstorm plug-in ^42^. In brief, SPRiNT builds on the specparam algorithm ^14^ and applies a short-time Fourier transform to decompose neural time series into aperiodic and periodic components within a sliding window. We used the following setting parameters: window length for short time fourier transform = 60 sec; overlap = 90%; frequency range 1-40 Hz; averaged FFT per time point = 1; while for the rest we used the default parameters provided by toolbox. SPRiNT thus yielded time series of both aperiodic (i.e., 1/f exponent) and periodic signals (i.e., oscillatory power), resulting in 120 values per ROI in the pre-ictal period and 66 in the resting state. SPRiNT results provided a time-frequency map for each ROI, allowing to investigate dynamic changes of the power spectrum. We grouped oscillatory results into results into the 5 canonical frequency bands: Delta (2-4), Theta (5-7), Alpha (8-12), Beta (13-20), Gamma (21-40).

### Inward/outward functional connections

Effective functional connectivity was computed using spectral Granger Causality (GC) via fieldtrip toolbox ^43^ with a model order of 10. To track dynamical changes of effective connectivity we used a sliding window approach with the same frequency range, window length and overlap of SPRiNT in order to obtain the exact same time resolution of dynamic GC and 1/f exponent. We then grouped GC results into the 5 canonical frequency bands previously described.

### Neurotransmitters maps

Neurotransmitter maps were extracted by the seminal work of Hansen and colleagues ^27^. Based on positron emission tomography (PET) with different ligands with a cohort of more than 1000 subjects, they openly provide whole-brain volumetric neurotransmitter density maps. We used the receptor density maps that were parcellated to 68 brain regions of the Desikan-Killiany atlas (https://github.com/netneurolab/hansen_receptors/tree/main/data/PET_parcellated/scale033). Specifically, we extracted the receptor density maps of the following neurotransmitters, as mainly related to E/I balance and seizure generation: GABA (GABAa) ^44^, acetylcholine (VAChT, M1, a4b2) ^45–50^, glutamate (NMDA, mGluR5) ^51–53^, opioid (MOR) ^54,55^ and serotonin (5-HTT, 5-HT1a, 5-HT2a, 5-HT1b, 5-HT4, 5-HT6) ^56–59^.

### Statistical analysis

For each time series, namely aperiodic exponent and power spectrum density both in pre-ictal and resting state condition, we applied a linear regression approach to extract regression slopes. To test the differences between slopes we applied two sets of contrasts: within conditions (Pre-Ictal and RS) contrasting Epileptogenic vs. Non-Epileptogenic Regions; between conditions: contrasting Epileptogenic in Pre-Ical vs. RS and Non-Epileptogenic in Pre-iCtal vs. RS. Specifically, from each regression we extracted the slope coefficients (*β*) and coefficient standard error (*SE*). To compare the difference of the two slopes we applied the following formula: 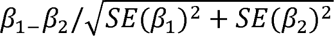. The *p*-values were extracted from the t-distribution.

Successively, correlations between aperiodic exponent and GC-derived inward and outward connectivity measures, as well as between aperiodic exponent and cortical thickness were performed using Spearman’s correlation with false discovery rate (FDR) correction for multiple comparisons.

Finally, a spin-permutation approach was applied to investigate the relationship between the cortical distribution of pre-ictal aperiodic exponent and the neurotransmitter density maps while controlling for the effects of spatial autocorrelation ^60^.

## Results

### E/I balance dynamics preceding seizure onset

Whole-brain dynamics are characterized by an increase in the aperiodic exponent as the seizure onset approaches, suggesting a global shift in the excitation/inhibition (E/I) balance toward inhibition (Figure 2A). Preictal slope of the aperiodic component was steeper as compared to the interictal period, for both the epileptogenic and non-epileptogenic regions (Epileptogenic regions pre-ictal v. RS: *t* = 2.032; *p* =.046; Non-Epileptogenic regions pre-ictal v. RS: *t* = 6.94; *p =* < .001). Notably, no significant slope differences were observed between epileptogenic and non-epileptogenic regions during the pre-ictal phase (*t* = 1.38; *p =*.16; see Fig. 2A), suggesting that E/I dynamic changes extend beyond the epileptogenic network.

**Figure 1.**
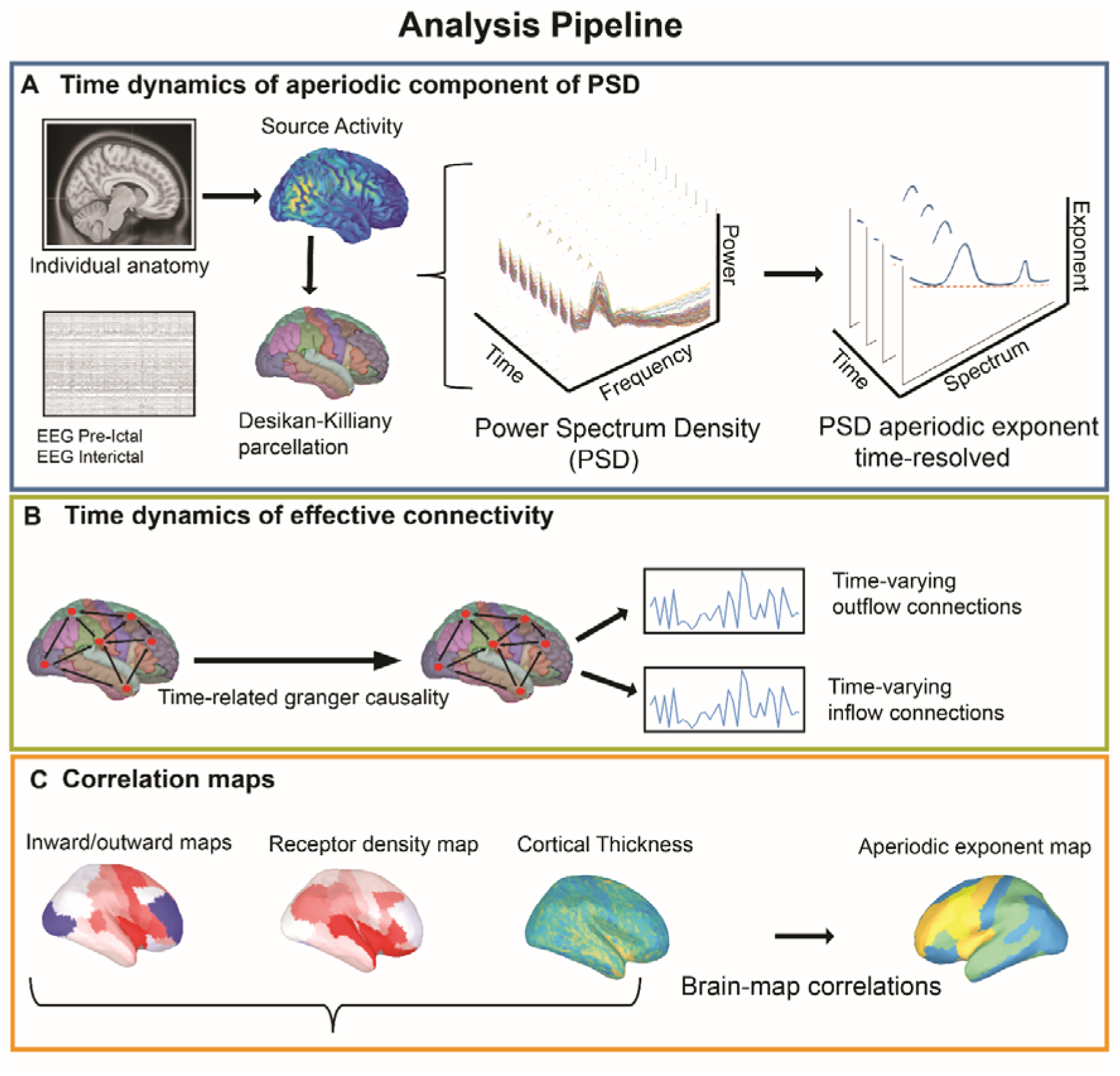
Analytical pipeline. (**A**) Electrical source imaging of the pre-ictal and interictal data; Desikan-Killiany atlas activity downsample and computation of dynamic changes of aperiodic component across time. (**B**) Time-dynamic of effective connectivity (spectral Granger Causality) to successively obtain time-varying inflow and outflow connections. (**C**) Correlation between the exponent of spectral aperiodic component with: in/outward connections, receptor density maps and cortical thickness.

**Figure 2.**
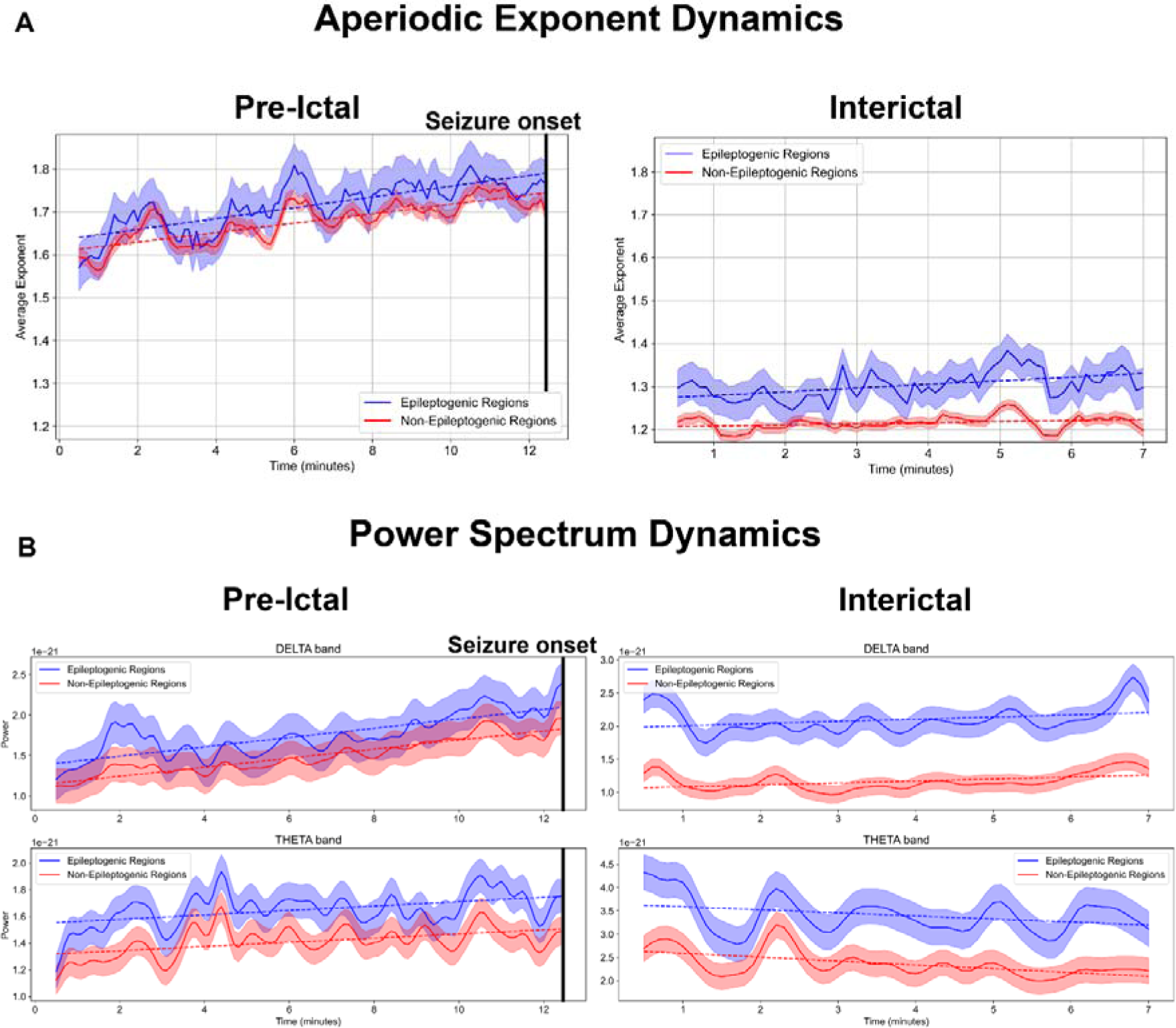
Aperiodic and periodic dynamics approach a seizure and in resting-state. (**A**) Dynamic changes in the aperiodic exponent. Top-left panel, an increase in the aperiodic exponent can be observed as the seizure approaches. Conversely, the top-right panel shows the aperiodic exponent during the interictal state, where no clear trend is detectable. (**B**) shows the temporal dynamics of delta and theta power (periodic component for pre-ictal and interictal conditions. Delta and theta increase as the seizure approaches. For both panels, blue lines indicate the behavior in epileptogenic regions, and red lines correspond to non-epileptogenic regions. The shaded area around the lines represents the standard error. The black line marks the seizure onset.

Similar findings were observed for the periodic component (power spectrum). Specifically, delta and theta frequency bands were characterized by a power increase as the seizure approached (see Fig. 2B). This result was supported by the difference in the slope of the PSD in time, when comparing pre-ictal to an interictal (resting state) time intervals (see Table 2). No differences were identified when comparing epileptogenic vs. non-epileptogenic slopes in delta and in theta bands (p > .05).

**Table 2.**
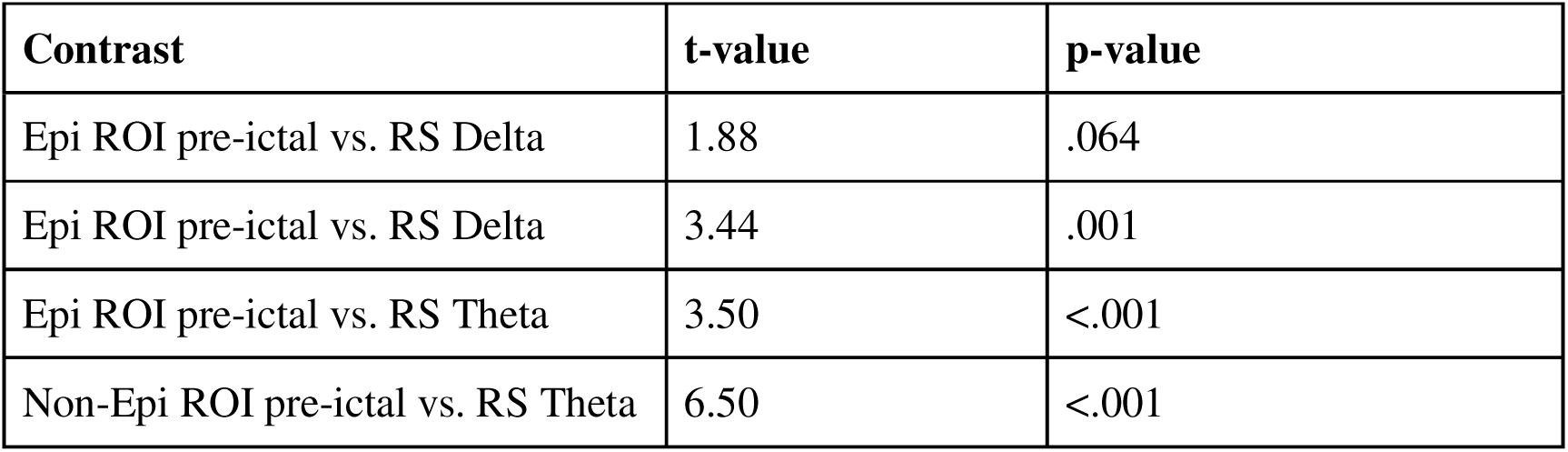
Slope analysis results. The table show the significant contrast of the slope analysis contrasting the dynamical changes in oscillatory activity (i.e. delta and theta bands).

### Inward/outward connections and relationship with exponent

When investigating time-dependent changes of in/outward connections in delta and theta bands in the preictal phase, we did not find any significant linear increase or decrease as the seizure approached (see Fig. 3A). Consequently, we shifted our focus to time-averaged inward and outward connections. Specifically, we averaged over time inward and outward connections of the pre-ictal period (13 minutes prior to seizure), separately. The same procedure was performed for the interictal period. We then compared connectivity profiles between epileptogenic and non-epileptogenic regions within pre-ictal and interictal phases, separately. Successively, in order to examine the relationship between E/I balance and functional organization (in/outward connectivity), we correlated time averaged aperiodic exponent (pre-ictal and interictal) with directed connectivity previously averaged over time.

**Figure 3.**
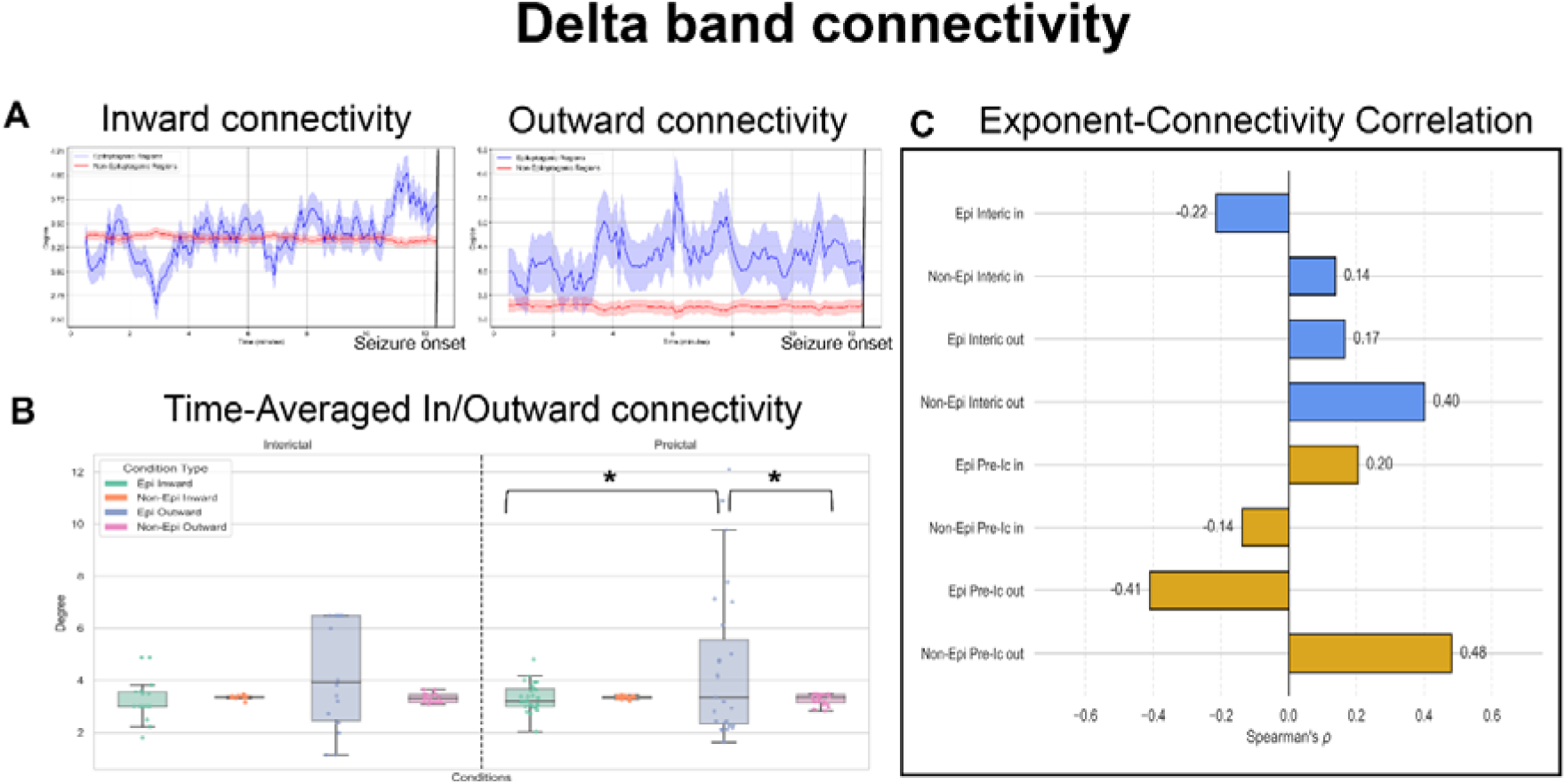
Effective connectivity in the delta band and its relationship with the aperiodic exponent. (**A**) Dynamics of the inward and outward number of connections (degree) in the delta band, for epileptogenic (blue lines) and non-epileptogenic (red lines) regions. Shaded areas indicate the standard error, and black vertical lines mark the seizure onset. (**B**) Distribution of the time-averaged degree during interictal and pre-ictal conditions using box plots. Significant comparisons are indicated by black lines with asterisks. (**C**) Spearman’s rho values for the correlation between the aperiodic exponent and the delta band inward (in) and outward (out) connectivity, represented as a barplot. Results are shown separately for epileptogenic (Epi) and non-epileptogenic (Non-Epi) regions, as well as for interictal (Interic) and pre-ictal (Pre-Ic) intervals.

Granger causality analysis in the delta band during the pre-ictal phase revealed a significantly greater number of outward connections compared to inward connections within epileptogenic regions (*t* = 2.12, *p* = .042). Furthermore, epileptogenic regions exhibited a greater number of outward connections compared to non-epileptogenic regions (*t* = 2.29; *p* = .030) (see Fig.3 B). Similar results were found for the theta band, and are reported in the Supplementary Materials (SM) (see Supplementary Fig.1A-B). By contrast, no significant differences were detected when comparing inward vs. outward connectivity within and between epileptogenic and non-epileptogenic regions.

As per the relationship between E/I and effective connectivity, larger inhibition (i.e., higher exponent values) were related to an increased number of outward connections in the non-epileptogenic region in the pre-ictal phase (rho = 0.48; *p* = .044). Interestingly, in the pre-ictal period, epileptogenic regions displayed an opposite correlation pattern as compared to the non-epileptogenic regions, with more outflow connections as the E/I balance shifted toward excitation. Even though the latter correlation does not survive multiple comparison correction (rho = −0.41; *p* = .067), it is worth mentioning the pattern inversion in the granger-exponent correlation between epileptogenic and non-epileptogenic regions in the pre-ictal period. On the other hand, both epileptogenic and non-epileptogenic, in the interictal phase, display the same correlation pattern between outflow connections and the aperiodic exponent, with no significant effect surviving multiple comparisons correction (see Fig.3C). Similar behavior was detected in the results related to the theta band (see Supplementary Fig.1C).

### Exponent-cortical thickness/receptors correlation

To investigate dynamic changes of structure-function coupling approaching the seizure, we performed a correlation analysis between cortical thickness and aperiodic exponent in the pre-ictal interval, in each time point, separately for epileptogenic and non-epileptogenic regions. We observed a stable correlation over time, suggesting non-significant changes in the structure-function coupling as the seizure approached (see Fig 4A). For this reason, we focused on the correlation between time-averaged value of the exponent in the pre-ictal period and the cortical thickness.

**Figure 4.**
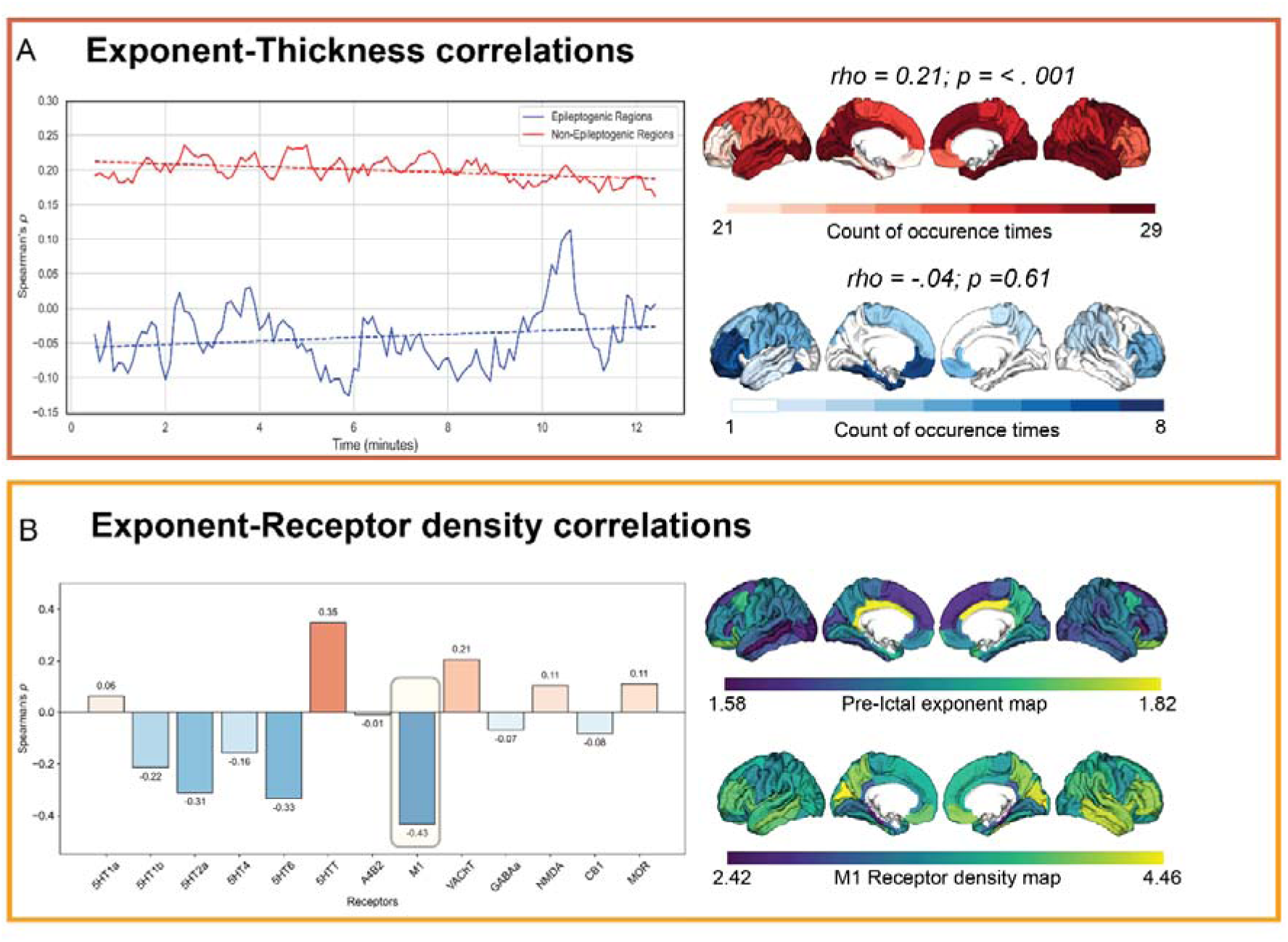
Correlational results. (**A**) In the top-left panel we show time-dependent connectivity between pre-ictal aperiodic exponent and the cortical thickness for epileptogenic (blu line) and non-epileptogenic (red line) regions. Brain plots on the top-right represent the counts (number of subjects) in which that specific region is considered non-epileptogenic (red regions) or epileptogenic (blue regions). (**B**) Spearman’s rho derived from spin-permutations between receptor density of multiple neurotransmitters and the brain map of the exponent of spectral aperiodic component averaged over time. The yellow shaded area highlights the correlation which survived FDR correction. Spatial distribution on the cortex (brain map) of the aperiodic exponent and the muscarinic receptor density are represented in the lower-right panel.

We identified a distinctive pattern between epileptogenic and non-epileptogenic regions. Specifically, larger values of cortical thickness were associated with increased inhibition (i.e., larger exponent values) (rho = 0.21; *p* = < .001), whereas no correlation was detected in epileptogenic regions (rho = −.04; *p* = 0.61; see Fig.4A).

Finally, E/I balance (i.e. aperiodic exponent) in the pre-ictal period was significantly negatively correlated with muscarinic receptor density (rho = -.433; *p* =.013; see Fig.4B). This suggests that an E/I balance is shifted towards inhibition in regions with lower muscarinic receptor density.

## Discussion

This study provides novel insight into the dynamic modulation of excitation/inhibition (E/I) balance in the human cortex preceding seizure onset, with a particular focus on its relationship to large-scale network interactions, cortical structure, and neurotransmitter receptor density. The findings suggest that seizure initiation may be preceded by a global, rather than strictly focal, reorganization of cortical excitability.

### Global Inhibitory Shift Precedes Seizure Onset

In the present work, by leveraging electrical source imaging, we showed that the aperiodic exponent, a non-invasive marker of E/I balance, dynamically increases as the seizure approaches. This finding is robust across both epileptogenic and non-epileptogenic regions, suggesting a global (whole brain) shift towards inhibition. Moreover, by comparing pre-ictal activity with an interictal condition, we showed the specificity of this dynamic pattern for the pre-ictal period.

Our results align well with recent intracranial EEG evidence suggesting a progressive inhibition in circadian rhythms of aperiodic exponent when approaching the seizure ^26^. Our results confirm the previous findings, and significantly enlarge the knowledge and understanding of this process. Firstly, contrary to SEEG, our approach allows whole-brain sampling, and is not limited to the investigation of regions already known to be involved in seizure generation. Secondly, thanks to the implementation of a comprehensive approach including structural and receptor mapping, we are able to infer the mechanisms which support our results.

As previously mentioned, the hypotheses that we tested here were generated based on the interictal suppression framework—mainly tested with intracranial recording—posing that the epileptogenic regions receive net inward inhibitory connections as a result of protective mechanisms, in the interictal phase ^19,20^. This is counterintuitive based on the knowledge that on a cellular level converging evidence linked seizures to an hyperexcitable state ^61,62^. In this light, the majority of current antiseizure medications (ASMs) work to restore E/I balance by either diminishing excitation or augmenting inhibition ^63^. However, recent theoretical expansions are enlarging our understanding of the link between E/I balance and seizure generation, suggesting that hyperexcitability may not be a universal driver for seizure occurrence ^10^. For example, dynamic assessment of the excitation/inhibition (E/I) balance in the rat hippocampus demonstrated that the initiation phase of seizure-like events (SLEs) is characterized by predominant inhibition in pyramidal neurons, with maximal synaptic excitability emerging during the ictal core of the SLE ^12^.This temporal dissociation suggests that early inhibitory dominance may constitute a network-level homeostatic response. Accordingly, we propose that the global inhibitory shift observed in EEG reflects large-scale cortical dynamics aimed at maintaining stability by actively steering the system away from the bifurcation threshold leading to seizure onset. This global modulation may serve as a last-line compensatory mechanism to prevent seizure onset, which ultimately fails as the ictal threshold is surpassed.

Moreover, the concurrent increase in low-frequency (delta and theta) oscillatory power in the pre-ictal phase aligns with previous observations of slowing cortical activity before seizures ^64,65^. Previous work has demonstrated that scalp EEG slow activity detected before the seizure may reflect underlying epileptiform activity, detected with SEEG ^66,67^. This synchronous increase in both aperiodic and periodic components strengthens the case for widespread changes in network excitability and suggests that these spectral signatures may be functionally coupled.

### Directionality of Functional Connectivity Differentiates Epileptogenic and Non-Epileptogenic Regions

The effective connectivity analysis may help to better elucidate whole-brain organization prior to a seizure, and its relationship with the E/I balance. First, in line with previous evidence, our findings reveal that epileptogenic regions are characterized by a larger number of outflow connections as compared to the inward connections. Additionally, outward connections number of epileptogenic regions is larger as compared to non-epileptogenic ones ^68^. These results align with previous intracranial findings, supporting the perturbatory role on the whole-brain network balance of the epileptogenic network ^65^. We observed a positive relationship between E/I balance and outward effective connectivity in non-epileptogenic regions. This association is specific to the period preceding seizure onset as opposed to the interictal period. These results align with the protective inhibitory framework, suggesting that the brain is trying to isolate perturbatory nodes, by reducing local connectivity and shifting the system E/I balance throughout inhibitory connections ^19,20,69^.

This compensatory mechanism is mainly expressed in the slow frequencies, being potentially linked to network tuning processes at whole-brain scale. Low-frequency brain rhythms play a crucial role in coordinating neural activity across widely distributed brain regions, facilitating whole-brain network communication. Their long wavelengths allow them to propagate effectively over large distances, enabling the coordination of brain activity in spatially separated areas ^70–72^.

Finally, the absence of consistent dynamic changes in connectivity leading up to seizures, along with more pronounced effects in time-averaged measures, suggests that structural organization constraints and baseline functional architecture may dominate over transient changes in driving seizure onset.

### Structural and Molecular Correlates of Pre-Ictal E/I Balance

In assessing the anatomical and neurochemical underpinnings of E/I dynamics, we found that cortical thickness was positively correlated with the aperiodic exponent in non-epileptogenic areas. This is in line with literature linking thicker cortex to stronger inhibitory control, possibly due to a greater abundance of inhibitory interneurons or supporting glial architecture ^73–75^. Interestingly, this structure-function coupling was absent in epileptogenic regions, which may reflect local pathological remodeling or dysmaturation that disrupts normative cortical organization ^76–78^.

Furthermore, spatial correlations with neurotransmitter receptor density highlighted a possible role of muscarinic cholinergic systems in shaping pre-ictal E/I dynamics. Regions with lower muscarinic receptor density were associated with greater inhibitory shifts, potentially implicating cholinergic tone in the loss of cortical excitability balance prior to seizures. All in all, this result aligns with the important role of the cholinergic system in seizure generation ^49,79,80^. This finding supports emerging evidence that neuromodulatory systems critically influence seizure susceptibility and offers a new avenue for targeted pharmacological interventions.

### Potential applications and limitations

One of the most promising outcomes of this work lies in the translational potential of dynamic E/I balance as a biomarker. The spectral aperiodic exponent offers a computationally efficient and non-invasive index of E/I balance that is both spatially and temporally sensitive ^14,42^. Given its consistent increase preceding seizures and its specificity this feature may serve as a critical predictive marker. In fact, the aperiodic exponent can be continuously tracked using scalp EEG, representing a sensitive feature with potential application in seizure prediction. Future development of closed-loop systems could leverage this metric for early seizure detection, potentially enabling adaptive neuromodulation to prevent seizures before clinical onset. While our findings are compelling, there are some limitations to mention. The study used a modest-sized retrospective cohort, with heterogeneity in terms of seizure characteristics. Furthermore, the correlative nature of the structure-function analyses could be better investigated with computation modelling ^24,81^. Nevertheless, the convergence of dynamic, anatomical, and neurochemical data provides a robust framework for future studies. Ongoing work should aim to replicate these findings in larger cohorts and evaluate their predictive value in prospective seizure forecasting pipelines. Integration with multimodal imaging and real-time EEG analytics may enable the development of personalized seizure prediction tools grounded in E/I balance dynamics.

### Conclusions

In summary, this study identifies a temporally dynamic and spatially widespread shift toward cortical inhibition at the whole-brain scale in the moments preceding seizure onset. These changes are intricately linked with the directionality of functional connectivity, regional structural features, and neurotransmitter receptor architecture. Together, our findings underscore the importance of considering both local and global neural dynamics in understanding and ultimately treating focal epilepsy.

## Data availability

The data that support the findings of this study are available on request to the corresponding author. The raw data are not publicly available due to privacy or ethical restrictions. All the scripts are available at the following GitHub page: https://github.com/simone-cuozzo/dynamic-fooof-prior-seizure.

