## Supplementary Material for "Dynamic excitation/inhibition balance preceding seizure onset and its link to functional and structural brain architecture"

**Inward/outward connections and relationship with exponent - Theta band**

No significant linear increase or decrease were found as the seizure approaches analyzing theta band changes of in/outward connections in the preictal phase (see Fig. 5A).

For the analyses on the time-averaged inward and outward connections the following pipeline was used: we averaged separately over time inward and outward connections of the pre-ictal and interictal periods and then compared connectivity profiles between epileptogenic and non-epileptogenic regions. In order to examine the relationship between E/I balance and functional organization (in/outward connectivity), we correlated time averaged aperiodic exponent (pre-ictal and interictal) with directed connectivity previously averaged over time. Similarly to the delta band results, in the theta band during the pre-ictal phase epileptogenic regions exhibited a greater number of outward connections compared to non-epileptogenic regions ($t$ = 2.16; $p$ = .040)(see Fig.5 B). The same result was found also in the interictal phase ($t$ = 2.42; $p$ = .026)(see Fig.5 B).

Finally, as per the relationship between E/I and effective connectivity, the pattern already found in the delta band was present also in theta band. Larger inhibition was related to an increased number of outward connections in the non-epileptogenic region in the pre-ictal phase (rho = 0.49; $p$ = .036). Conversely, in the pre-ictal period epileptogenic regions displayed an opposite correlation pattern as compared to the non-epileptogenic regions, with more outflow connections as the E/I balance shifted toward excitation even though the latter correlation does not survive multiple comparison correction (rho = -0.42; $p$ = .055) (see Supplementary Fig.1)

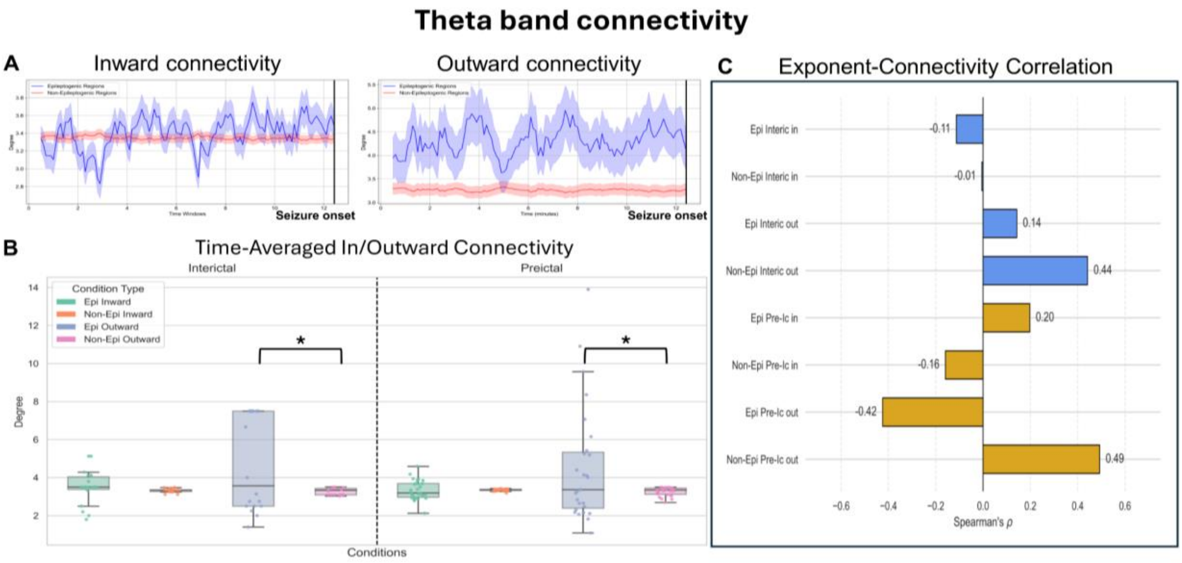


**Supplementary Figure 1 Effective connectivity in theta band and its relationship with the aperiodic exponent.** (**A**) Dynamics of the inward and outward number of connections (degree) in the theta band, for epileptogenic (blue lines) and non-epileptogenic (red lines) regions. Shaded areas indicate the standard error, and black vertical lines mark the seizure onset. (**B**) Distribution of the time-averaged degree during interictal resting-state and pre-ictal conditions using box plots. Significant comparisons are indicated by black lines with asterisks. (**C**) Spearman's rho values for the correlation between the aperiodic exponent and the theta band inward (in) and outward (out) connectivity, represented as a barplot. Results are shown separately for epileptogenic (Epi) and non-epileptogenic (Non-Epi) regions, as well as for interictal (Interic) and pre-ictal (Pre-Ic) intervals
